# Antibiotic utilisation and the impact of antimicrobial resistance action plan on prescribing among older adults in New Zealand between 2005 and 2019

**DOI:** 10.1101/2023.02.15.23285986

**Authors:** Tichawona Chinzowu, Te-yuan Chyou, Sandipan Roy, Hamish Jamieson, Prasad S Nishtala

**Author notes:** **Address correspondence to:** Tichawona Chinzowu, Department of Pharmacy and Pharmacology, University of Bath, Bath, BA2 7AY, United Kingdom.

## Abstract

**Purpose:** The objectives of this study were to examine the overall utilisation of systematic antibiotics and the impact of the launch of the antimicrobial resistance action plan in older adults (65 years or more) living in New Zealand between 2005 and 2019, using data from a national database (Pharmaceutical collections), stratified by antibiotic class, patient age, and sex.

**Methods:** Population-level systemic antibiotic dispensing data for older adults in New Zealand was analysed using repeated cross-sectional analysis between 01/01/2005 and 31/12/2019. Data were extracted on the prescribed systemic antibiotics using a unique identifier for each case. Antibiotic utilisation was measured in DDD/TOPD values. In addition, an interrupted time series (ITS) analysis using the autoregressive integrated moving average (ARIMA) models was performed to determine the impact of the antimicrobial resistance action plan.

**Results:** Most of the antibiotic classes included in this study showed a significant overall decrease in utilisation ranging from 38.64% for sulphonamides to 80.64% for fluoroquinolones compared to 2005. Systemic antibiotic utilisation decreased by 49.6% from the predicted usage between January 2018 and December 2019 following the launch of the antimicrobial resistance action plan. ARIMA model supported the reduction in utilisation with a step change of -0.2206 and slope change of -0.0029.

**Conclusions:** The ITS analysis has demonstrated that the intervention may have hugely impacted the antibiotic utilisation rate among older adults. Further studies are required to determine whether the reduced consumption rates of antibiotics are associated with reduced rates of antibiotic-associated adverse drug events such as acute kidney injury and haematological abnormalities.

## Introduction

Inappropriate prescribing of antibiotics is an important driver of the rising global antibiotic resistance and a direct threat to the safety of patients due to the increased risk of adverse drug events such as acute kidney injury, haematologic, gastrointestinal, dermatologic and hepatobiliary abnormalities [1-3]. Outpatient and community utilisation of antibiotics is on the rise, with older adults (>65 years of age) receiving the highest prescribing rate compared to any other age group [4]. In the United States, older adults are one and half times more likely to receive antibiotics than younger adults in a year [5, 6]. In over 46% of visits involving non-bacterial respiratory tract infections, adults ≥65 receive unnecessary antibiotics, most of which are broad-spectrum agents [7, 8]. This increased rate of inappropriate prescribing raises concerns for older adults who are already at increased risk for serious adverse drug events due to age-related physiologic changes, comorbidities, and polypharmacy [9]. Antibiotics, as a class, are the third commonest cause of adverse drug events among older adults. Nearly 15% of all emergency department visits for antibiotic-associated adverse drug events occur in older adults [10].

Specific antibiotic classes pose increased safety risks for older adults. For example, fluoroquinolones have been reported to put older adults at an elevated risk of serious adverse events, including tendon rupture, blood sugar disturbances, peripheral neuropathy, delirium and aortic dissection [11]. Macrolides, commonly prescribed to older adults, were associated with an elevated risk of cardiac arrhythmia and death, particularly in males [12]. Cotrimoxazole requires close monitoring in older adults due to the risk of pre-existing renal insufficiency necessitating dose adjustment [1].

There was a significant decrease in annual antibiotic usage between 2014 and 2018 among most countries in the world Health Organisation (WHO) Europe region, including the United Kingdom, Germany, and Finland [13]. In their study, Andersson et al. [14] showed consistently higher antibiotic usage in Australia compared to Sweden. In New Zealand, systemic antibiotic consumption increased from 17.31 to 25.79 DDD per 1000 per day between 2006 and 2014 [15]. In another New Zealand study, systemic antibiotics’ consumption increased by 2% between 2010 and 2015 among older adults [16].

For population-level health interventions, the interrupted time series (ITS) analysis is a valuable tool for evaluating their effectiveness, provided they were implemented at a well-defined time point [17]. Segmented regression is a commonly used approach for ITS analysis. However, it is not adequate in autocorrelation and seasonality [18]. An alternative method, the ARIMA model, can accommodate both seasonality and autocorrelation [18].

Few studies describe the utilisation of systematic antibiotics at the population level, specifically among older adults in New Zealand, and how the utilisation rates respond to interventions primarily aimed at mitigating the development of antimicrobial resistance. Therefore, the objectives of this study were to examine the overall utilisation of systematic antibiotics and the impact of the launch of the antimicrobial resistance action plan in older adults (65 years or more) living in New Zealand between 2005 and 2019, using data from a national database (Pharms), stratified by therapeutic class, age, and sex, based on the World Health Organisation Collaborating Centre for Drug Statistics Methodology (WHOCC) Anatomical Therapeutic Chemical (ATC) classification.

## Methods

### Study design

Population-level systemic antibiotic dispensing data for older adults in New Zealand was analysed using repeated cross-sectional analysis between 1^st^ January 2005 and 31^st^ December 2019. Data were extracted on the prescribed systemic antibiotics using a unique identifier for each case and analysed for utilisation in DDDs according to patients’ antibiotic class, age, and sex. This study included dispensing claims data for all funded antibiotics by community pharmacies.

### Data source

From 1st January 2005 to 31st December 2019, de-identified individual dispensing claims data for older adults (65 years and above) were extracted from the Pharmaceutical Collections [19, 20]. Pharmaceutical collections are the New Zealand Ministry of Health’s national dispensing claims database which captures subsidised prescriptions dispensed by community pharmacies [21]. Antibiotics were categorised using the ATC developed by WHOCC. The following antibiotic classes were included in this study: penicillins (J01C), macrolides (J01F), cephalosporins (J01D), tetracyclines (J01A), sulphonamides (J01E), fluoroquinolones (J01M), aminoglycosides (J01G), and ‘other antibiotics’ (J01X) [22]. In addition, metronidazole, nitrofurantoin, and vancomycin were classified as ‘Other antibiotics’ (J01X).

### Data analysis

DDDs were computed for each antibiotic dispensed to an individual yearly and normalised by age group, antibiotic class, and sex at a population level. In this study, the DDD per 1000 older people per day (TOPD) measures the proportion of people being treated with a daily dose of antibiotic dispensed within a defined study area per 1000 older people per day. Antibiotic utilisation by class, sex and age were also measured in DDD/TOPD values. The DDDs were computed for each antibiotic utilised by an individual yearly, per annual population size of New Zealand between 2005 and 2019. The R statistical software (version 4.2.0) was used in all analyses [23].

### Interrupted time series analysis

Using the R statistical software (version 4.2.0), we determined the effect of introducing the antimicrobial resistance action plan in January 2018 in New Zealand on antibiotic prescribing and utilisation rates among older adults. We performed ITS analysis using the ARIMA model [18] on monthly antibiotic utilisation rates to understand the effect of the implemented national antimicrobial resistance action plan [18]. Monthly DDD/TOPDs were computed above for the study period and population. The pre-intervention period was from January 2005 to December 2017, and the post-intervention period was from January 2018 to December 2019.

Firstly, the time series data was plotted to understand any patterns, including trends and seasonal variations. Then, the forecast R package [23] was used to determine *p/P* and *q/Q* parameters. Next, stationarity was induced using the first-order difference where d = 1 and D = 1 for trend and seasonal differences, respectively. Next, we plotted the autocorrelation and partial autocorrelation functions (ACF/PACF) of stationarity data to view and confirm the ARIMA orders from the automated algorithms. Finally, the Ljung-Box test [18] was used to test whether residuals for the model were white noise or not.

Counterfactual values for the post-intervention period were determined and plotted together with observed values to visualise the impact of the intervention on the antimicrobial utilisation rate among older adults.

## Results

### Overall trend

Table 1 shows the percentage change in the utilisation of funded systemic antibiotics among older adults between 2005 and 2019 in New Zealand. Overall systemic antibiotic utilisation decreased by 56.28% between 2005 and 2019, averaging an annual decrease of 4.02% (Table 1). However, utilisation remained high from 2005 to 2017, declining between 2018 and 2019 (Figure 1) following the implementation of the New Zealand Antimicrobial Resistance Action Plan [24].

**Table 1:**
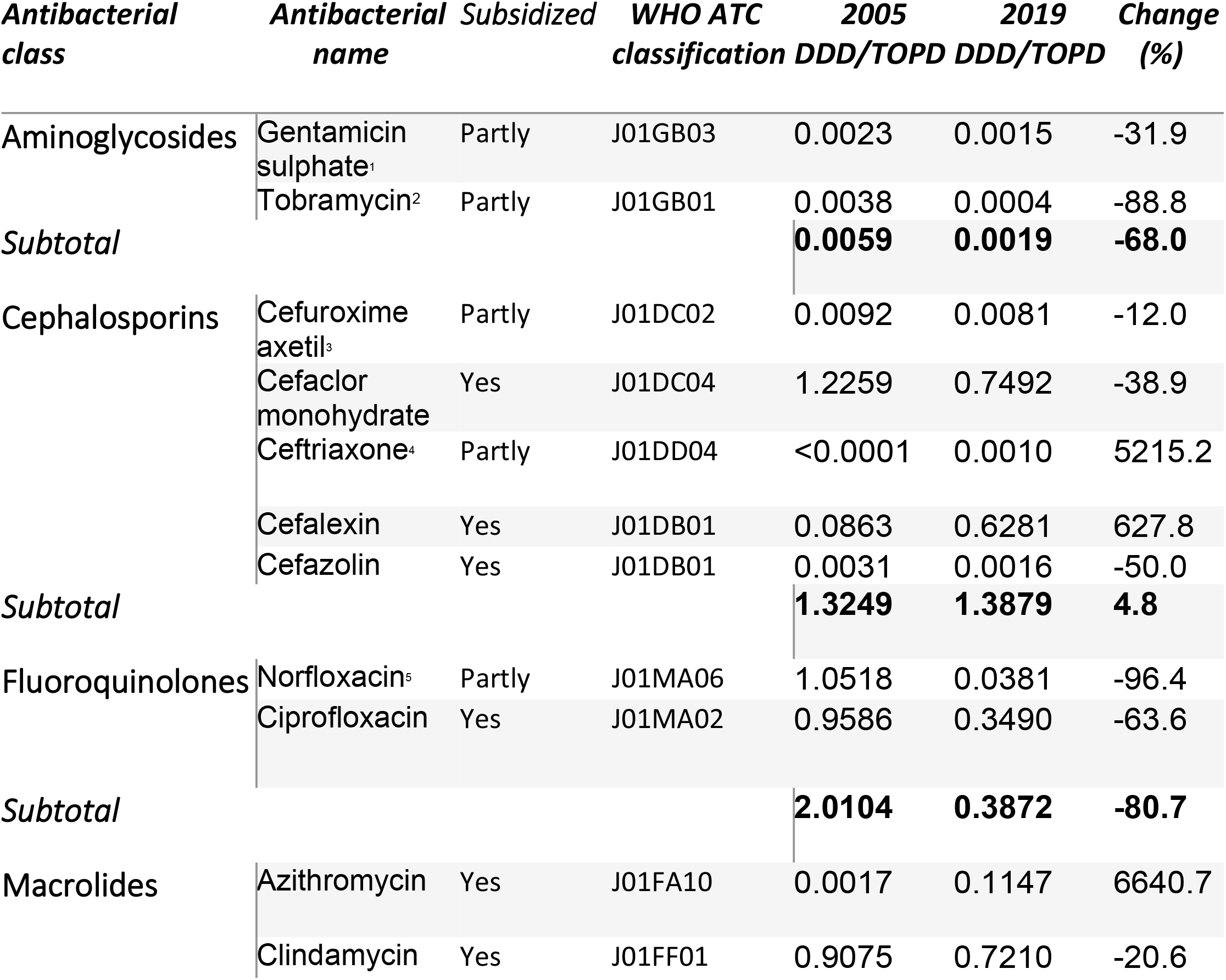
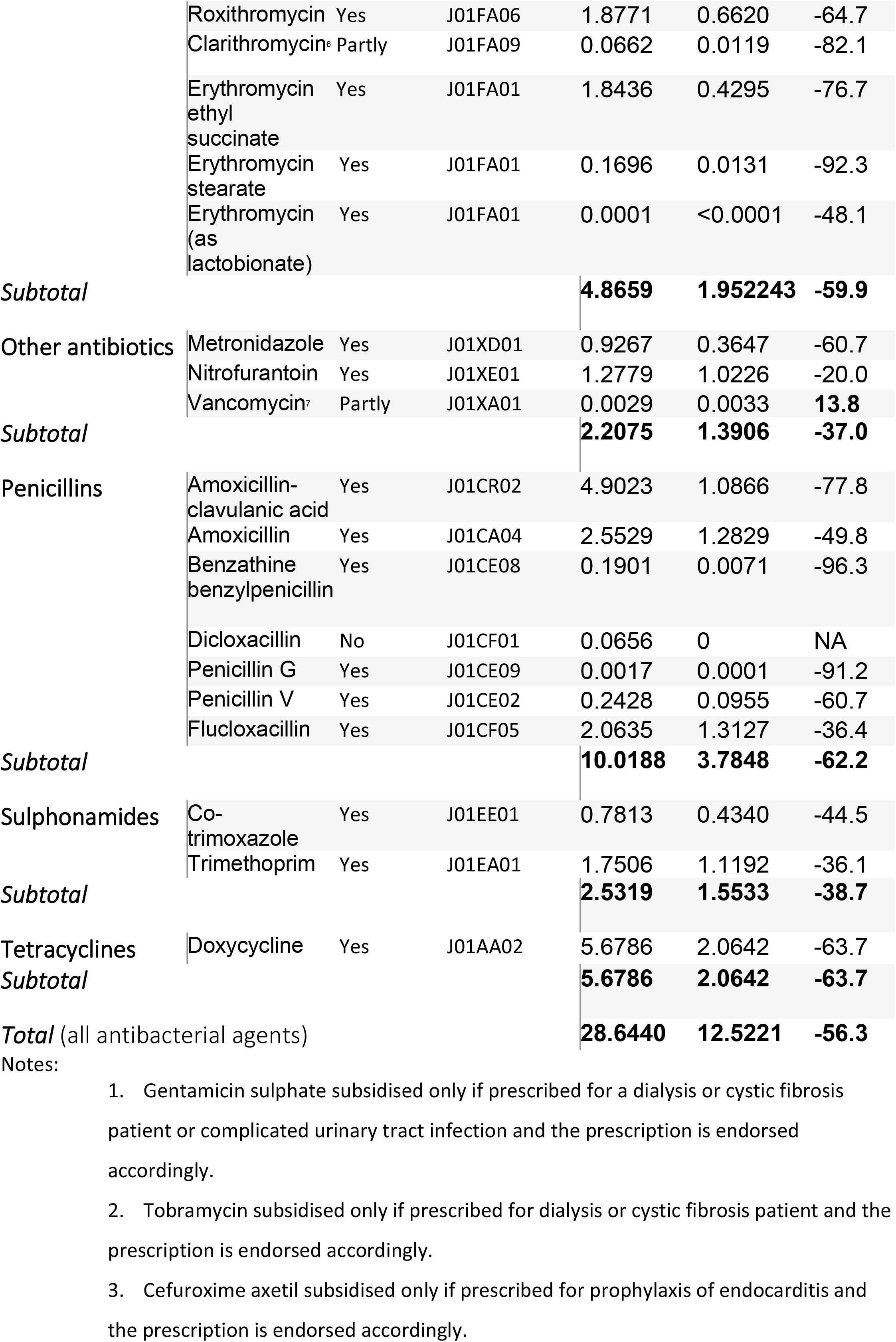
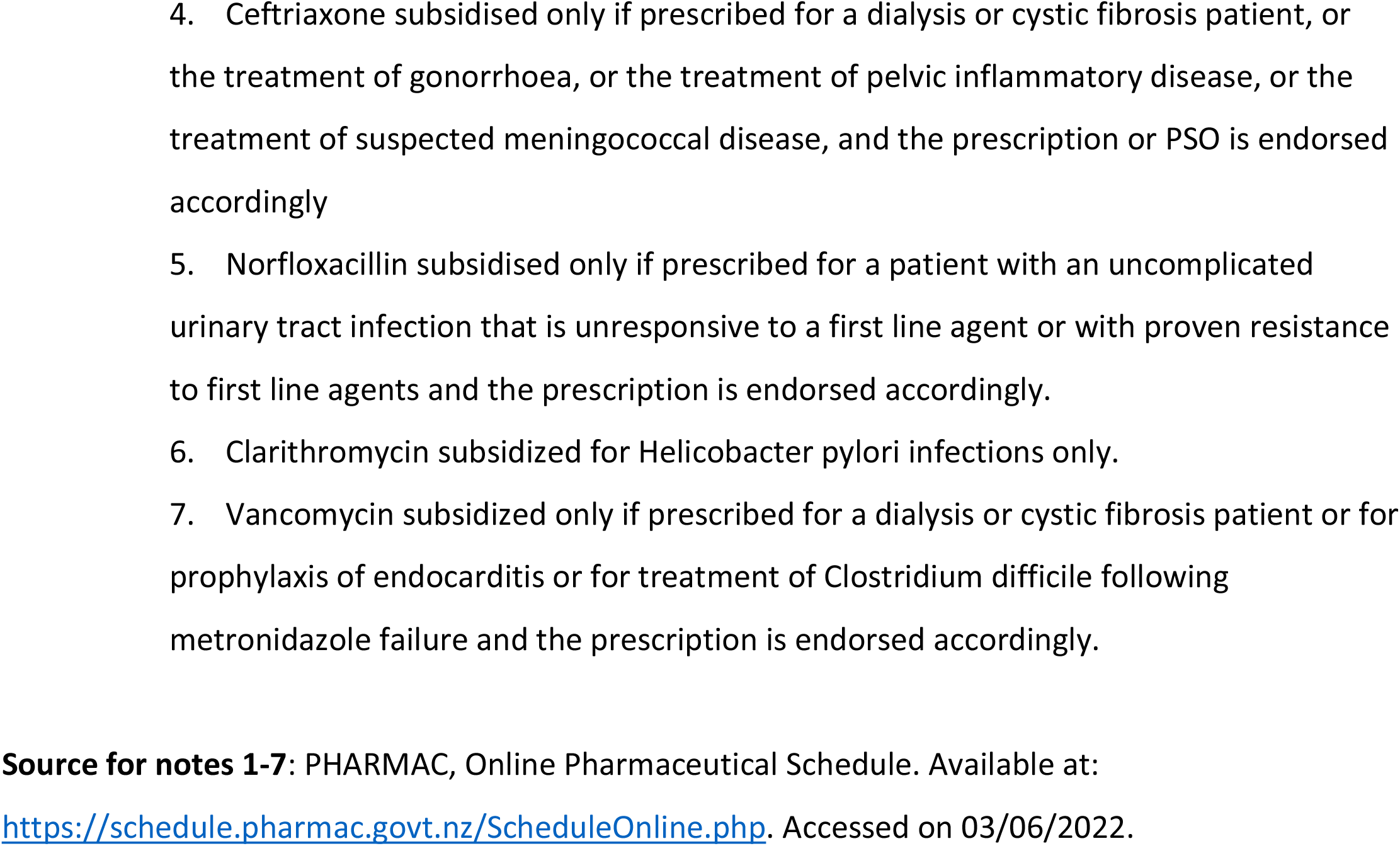
Utilization of antibacterial agents (in defined daily dose per 1000 older people per day [DDD/TOPD] values) between 2005 and 2019.

**Figure 1:**
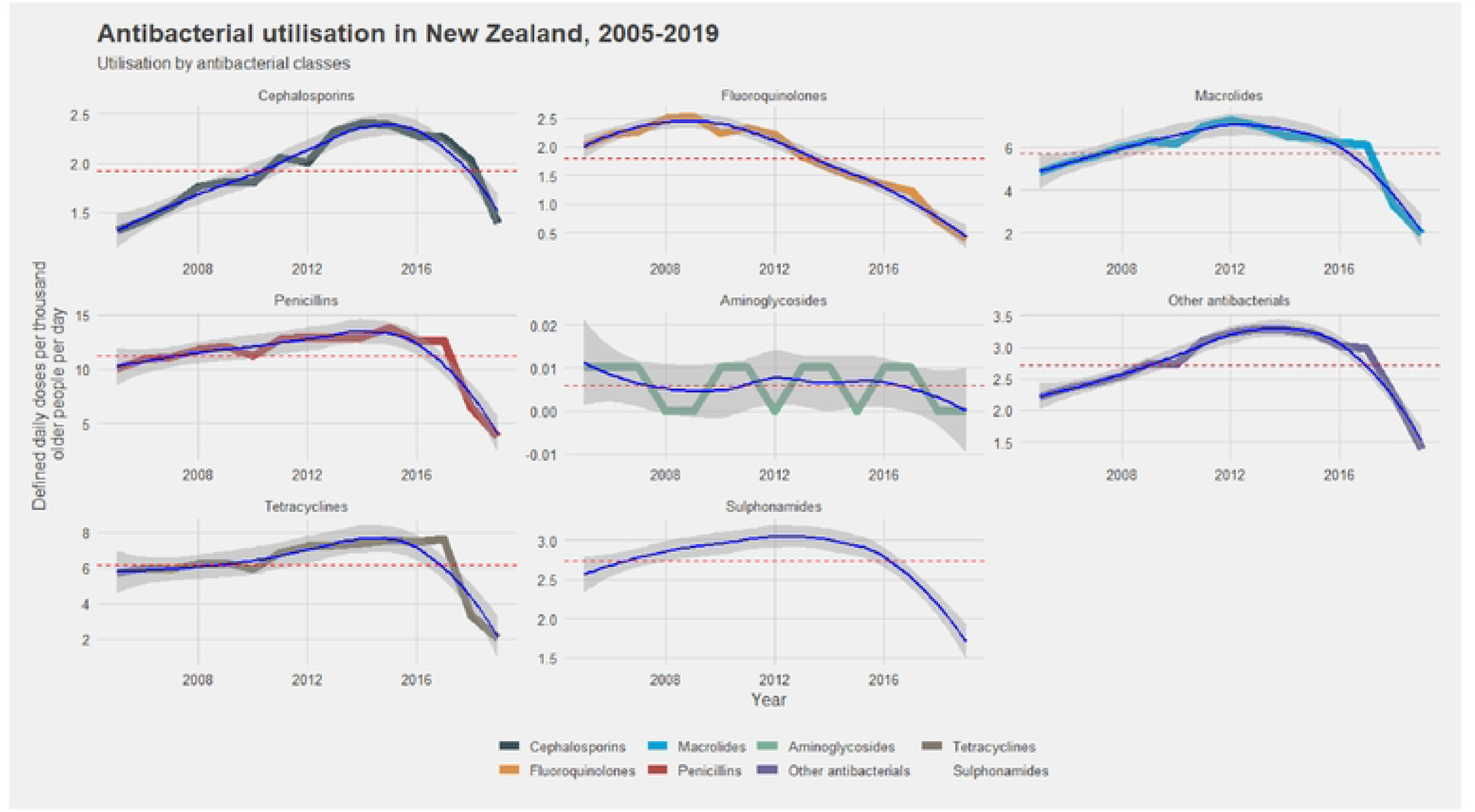
Systemic antibacterial utilisation by antibiotic class, expressed as defined daily doses per thousand older people per day (DDD/TOPD), from 2005 to 2019 for older adults (65 years or more) in New Zealand.

Most antibiotic classes included in this study showed a significant decrease in utilisation ranging from 38.64% for sulphonamides to 80.64% for fluoroquinolones. However, cephalosporins showed a 4.76% increase in usage during the study period, and this was largely attributed to the increased utilisation of cephalexin and ceftriaxone. Although macrolide utilisation decreased significantly, the utilisation of azithromycin increased by just over 66 times that of 2005 by the end of 2019. Vancomycin only showed an overall increase of 13.8% in utilisation during the study period. In 2019, the top four single most utilised antibiotics were doxycycline (DDD/TOPD = 2.06), flucloxacillin (DDD/TOPD = 1.31), amoxicillin (DDD/TOPD = 1.28), and trimethoprim (DDD/TOPD = 1.12) (Table 1).

### Utilisation by antibiotic classes

Except for aminoglycosides and fluoroquinolones, all included antibiotic classes showed a steady increase in utilisation until 2016 (Figure 1), followed by a sharp decline. Fluoroquinolone utilisation started declining in 2012, while aminoglycoside utilisation remained very low. The most utilised antibiotic classes were penicillins (DDD/TOPD = 3.78), tetracyclines (DDD/TOPD = 2.06), and macrolides (DDD/TOPD = 1.95) (Table 1). The biggest reduction in utilisation between 2005 and 2019 was on fluoroquinolones, which dropped by 80.7% (Table 1).

### Utilisation by antibiotic class and age

The total systemic antibiotic consumption increased with age but remained relatively steady from 2005 to 2017 (Figure 2). Penicillins were the most utilised antibiotics across all age groups. In addition, the DDD/TOPD for all classes, except aminoglycosides, increased with age.

**Figure 2:**
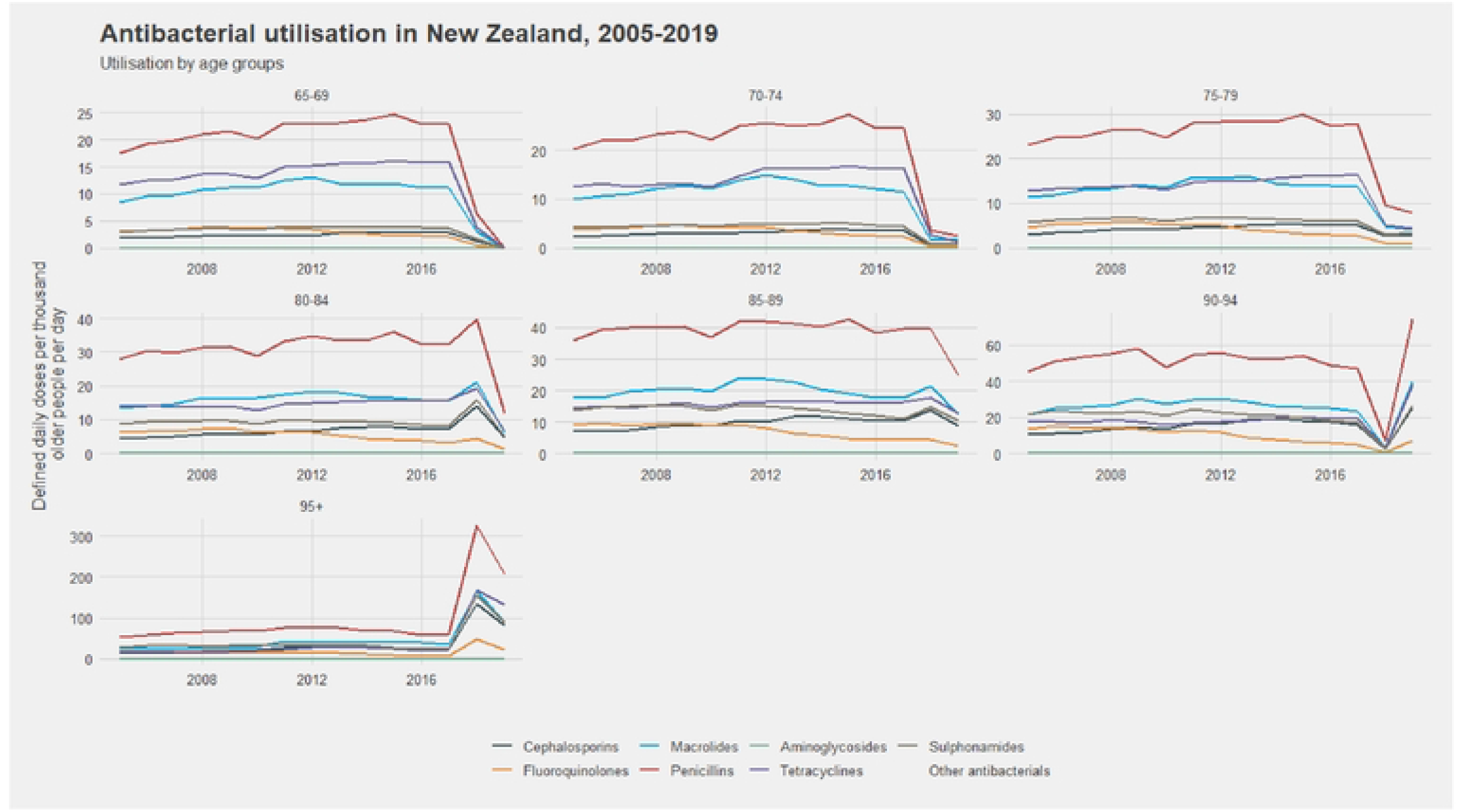
Systemic antibacterial class consumption by age group, expressed as defined daily doses per thousand older people per day (DDD/OPD), from 2005 to 2019 for older adults (65 years or more) in New Zealand

### Utilisation by antibiotic class, age, and sex

The rate of antibiotic consumption was constant in both males and females for all antibiotic classes (except fluoroquinolones) and age groups, except for the 95 years old and above males, whose consumption rate has been steadily increasing in all antibiotic classes (Figure 3). Between 65 and 79 years of age, antibiotic utilisation was similar for both sexes. Females had higher utilisation than males from age 80 to 94 years. Among those 95 years and above, males had greater utilisation than females. Fluoroquinolone utilisation has been decreasing in all age groups for both males and females as early as 2012, except among the 95 years and above males, where the decrease in consumption started in about 2015.

**Figure 3:**
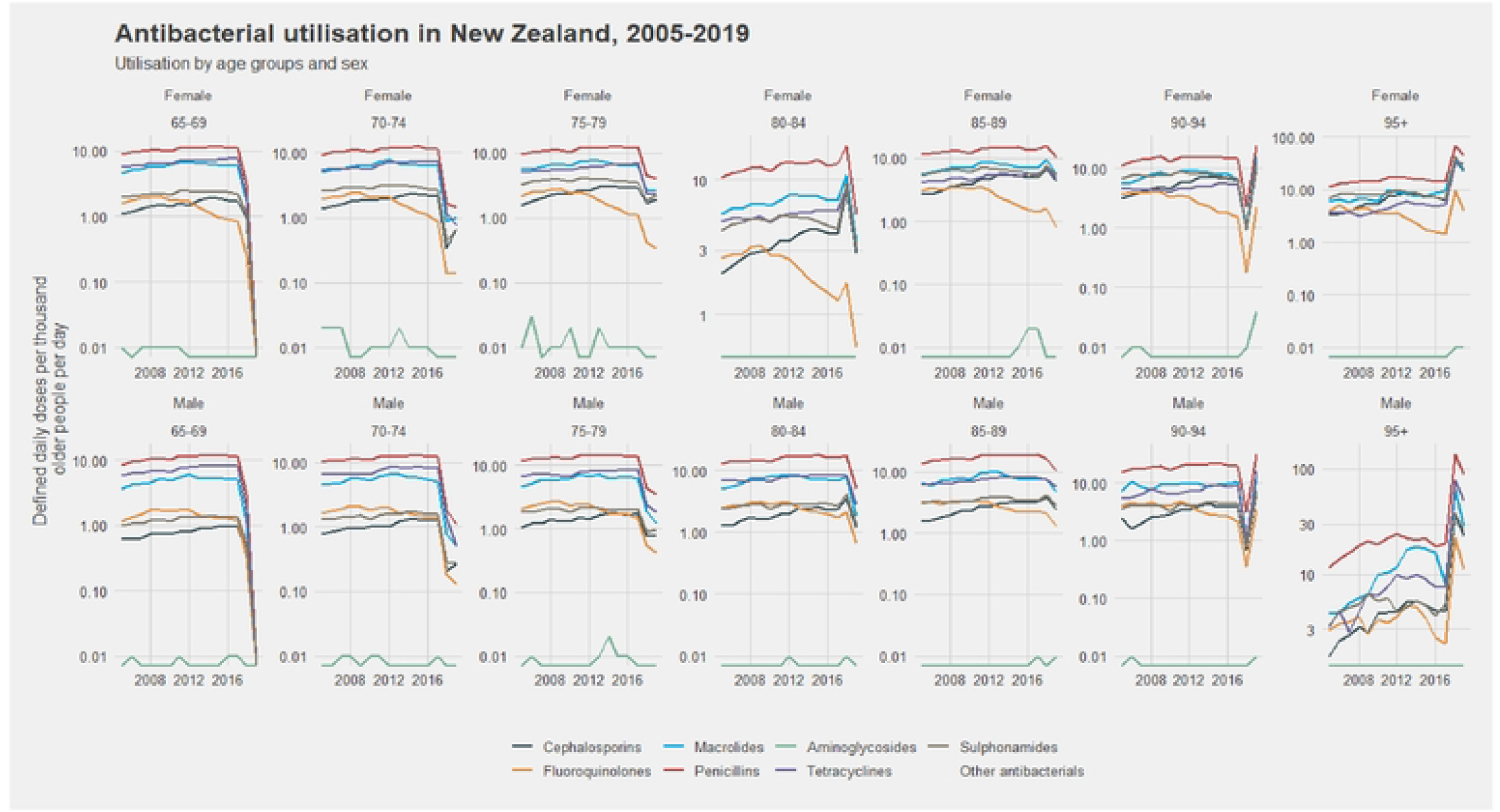
Systemic antibacterial class consumption by age group and sex, expressed as defined daily doses per thousand older people per day (DDD/TOPD), from 2005 to 2019 for older adults (65 years or more) in New Zealand.

### Interrupted time series analysis

The observed data showed a slight upward trend and possible seasonality on analysis (Supporting file, Figures S1, S2 and S3). Before the intervention, the visible trend and seasonality determined stationarity by first difference (d = 1) and seasonal difference (D = 1). According to the undifferenced and differenced ACF/PACF plots obtained, the former showed autocorrelation at lag 12 (Supporting file, figure S4 and S5). After running the automated algorithm in R, the best model selected was (3,1,0) (0,1,1) [12]. Thus, autocorrelation order (p) was 3, the moving average order (q) was 0, the autocorrelation order for the seasonal part (P) was 0, and the moving average order for the seasonal part (Q) was 1. The p-value for the Ljung-Box test for white noise was 0.51 at 36 lags (Spporting file, figure S6); therefore, the chosen model had a good fit. Figure 4 is the final ARIMA model showing predicted values in the absence of an antimicrobial resistance action plan (counterfactual) compared to observed values. The estimated step change was minus 0.2206 (95% CI: -0.4619 – 0.0207) and the estimated change in slope was -0.0029 (95% CI: -0.0097 – 0.0039).

**Figure 4:**
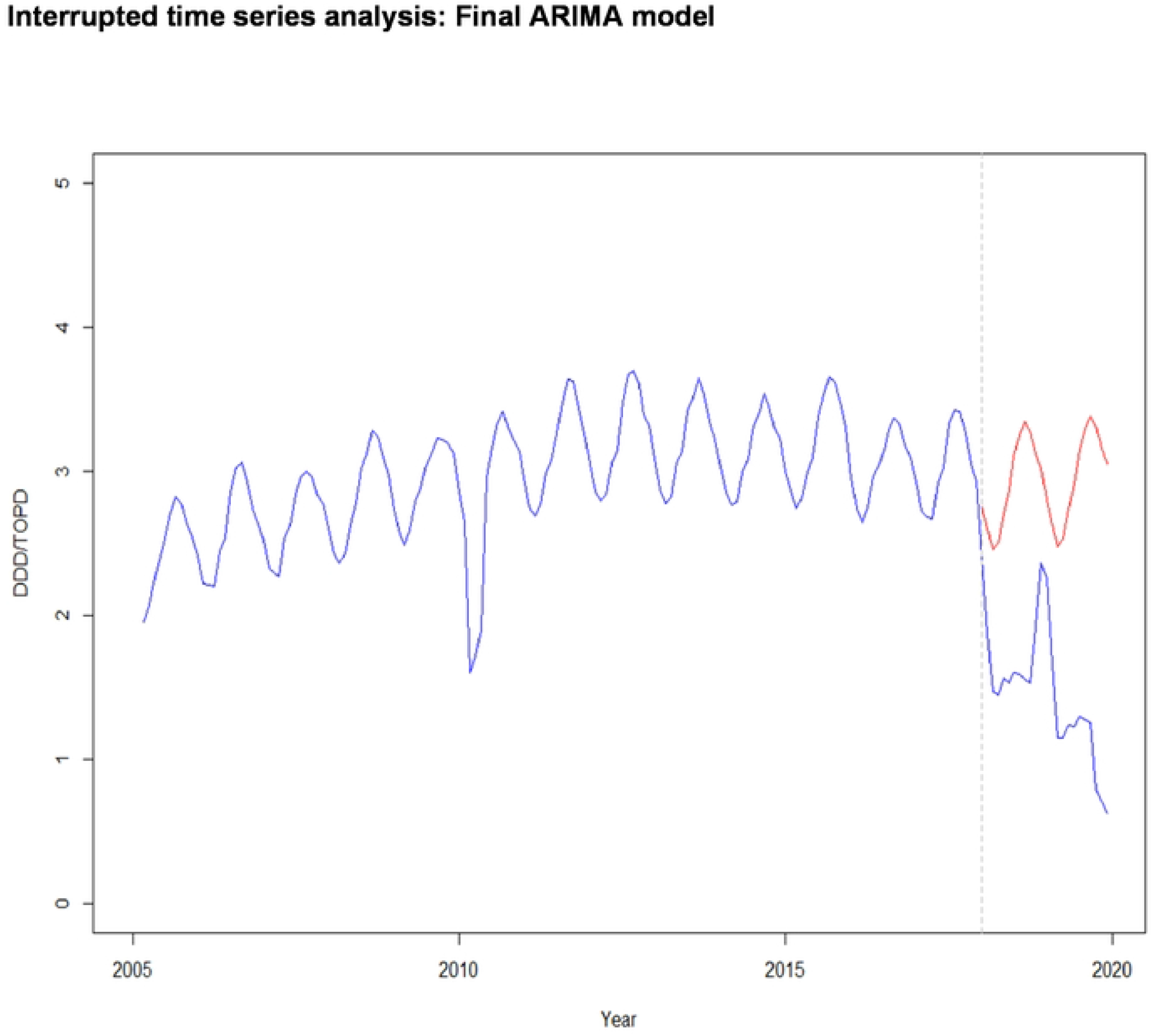
Final ARIMA model showing predicted values in the absence of antimicrobial resistance action plan (counterfactual) compared to observed values. Step = -0.2206 (95% Cl: -0.4619 - 0.0207). Ramp = -0.0029 (95% Cl: -0.0097 - 0.0039) Key: Counterfactual = Red. Observed values = Blue. Intervention date = Grey dotted vertical

## Discussion

### Overall trend

This study found that the overall utilisation of subsidised systemic antibiotics among older people in New Zealand dropped by 49.6% of the predicted utilisation rate for 2018/2019 following the implementation of the antimicrobial resistance action plan [24]. In a previous study, systemic antibiotic utilisation was reported to have increased 49% between 2006 and 2014 in New Zealand [15]. This increase is also reflected in this study, where utilisation of most antibiotic classes steadily increased until 2015 (Figure 1 and table 2). However, this was followed by a sharp decrease to more than 56% below the 2005 utilisation rate between 2018 and 2019.

We concluded that implementing the antimicrobial resistance action plan contributed significantly, but not exclusively, to the significant drop in total antibiotic utilisation rates in New Zealand in 2018 and 2019. In 2015, Thomas [25] called for immediate action to reduce antimicrobial prescribing in New Zealand, particularly in the community where most infections were self-limiting. His recommendations included not prescribing antibiotics where the infection was likely viral, not prescribing antibiotics for sore throat in all over eighteen (except for Māori and non-Pacific ethnicities), substituting amoxicillin-clavulanic acid with penicillin for Streptococcus pyogenes skin infections, and no fluoroquinolones for urinary tract infections. These recommendations were supported by the Antibiotic Consumption in New Zealand, 2006 – 2014 surveillance report [26]. Significant reduction in amoxicillin and amoxicillin-clavulanic acid utilisation rates would greatly impact total antibiotic utilisation since these two antibiotics contributed more than 42% of the total antibiotic utilisation rates for the period 2006 - 2014 [26]. The utilisation of these penicillins, and to a lesser extent macrolides and tetracyclines, followed a seasonal pattern with peaks during the winter months (June to August) and troughs during the summer months (December to February) in New Zealand [26]. The highest use of penicillins was reported among older adults of the Pacific Peoples, European (and other), Middle Eastern, Latin American, and African (MELAA) ethnicities. The utilisation of amoxicillin and amoxicillin-clavulanic acid doubled between 2006 and 2014, possibly overusing due to overlapping clinical features of indicated conditions with viral infections [26].

The antimicrobial resistance action plan was developed through a collaboration of various human, animal and agricultural stakeholders to respond to the ever-rising global challenge of antimicrobial resistance [24]. The implementation would be spread over five years from 2017, with the first year mainly for fact-finding, training, and resource mobilisation before rolling out the activities from year two onwards. One of the main objectives focussed on the optimisation of the use of antimicrobial medicines, whereby in 2017, the focus was on investigating inequalities in antimicrobial prescribing in community hospital settings and, from 2018, rolling out the appropriate activities to reduce antimicrobial consumption while promoting appropriate prescribing began [24]. In our study, we, therefore, considered January 2018 as the date of starting the measures recommended to reduce antimicrobial utilisation in New Zealand.

The ITS analysis, as shown in figure 4, has demonstrated that this intervention significantly impacted the antibiotic utilisation rate among older adults. The huge drop in total antibiotic utilisation rate was probably due to a huge drop in prescribing amoxicillin or amoxicillin-clavulanic acid following the implementation of the action plan. The Best Practice Advocacy Centre for New Zealand (BPACNZ) supported this and noted a sustained decrease in dispensed oral antibiotic prescriptions per 1000 registered patients [27]. In 2018 and 2019, there was an average of 3% to 5% annual drop in dispensed antibiotic prescriptions, according to the BPACNZ [27]. This decrease was attributed to New Zealand’s improved antimicrobial stewardship. Our findings were further supported by Thomas et al.[28], who concluded that there was an overall decrease in dispensing of antibiotics among adults 60 years or above in New Zealand between 2015 and 2018. In their study, Thomas et al.[28] found that community antibiotic dispensing, measured as defined daily doses per 1,000 inhabitants per day, decreased by 4.6% per year during 2015-2018, with large reductions in dispensing of amoxicillin-clavulanate [28].

In a recent study, overall antibiotic utilisation was consistently higher in Australia compared to Sweden between 2006 and 2018 [14]. In Australia, the overall utilisation was steady, ranging from 21.8 DDD/TOPD in 2006 to 22.2 DDD/TOPD in 2018, whereas Sweden experienced a 26.2% decrease in utilisation between 2006 and 2018. Compared to Australia and Sweden, our results showed that New Zealand’s drop in overall antibiotic utilisation was the fastest, which could have been necessitated by the government’s strong response to the increasing global challenge of antimicrobial resistance in 2016 [29].

The top four most utilised antibiotic agents in 2019 were doxycycline, flucloxacillin, amoxicillin, and trimethoprim (Table 1). This finding agrees with previous studies where doxycycline and amoxicillin consistently appeared on the top list of heavily consumed single antibiotics [14, 15]. However, in a separate US study, the top four most used antibiotics among older adults (65 years or more) were azithromycin, ciprofloxacin, amoxicillin, and cefalexin [30]. The differences in the usage of individual antibiotics are possibly due to different local and national antibiotic stewardship policies, treatment guidelines, subsidy arrangements, and non-adherence. [13, 31, 32].

### Utilisation by antibiotic class

This study showed that in 2019 the most utilised antibiotic classes were penicillins, tetracyclines, and macrolides (Table 1). Previous studies have also found penicillins, extended-spectrum penicillins and beta-lactamase sensitive penicillins as the most heavily consumed antibiotic classes in New Zealand, Australia, and Sweden, respectively [14, 15]. This general agreement in utilisation trends, by class, of systemic antibiotics may indicate an internationally improved antibiotic stewardship.

### Utilisation by age group and sex

In this study, antibiotic utilisation rates generally varied with age and sex, increasing in the older age group (Figure 2). Williamson et al. [15] also found that antibiotic utilisation rates increased with age among the New Zealand population and were significantly and consistently higher in females than males between 2006 and 2014. Our study showed that antibiotic utilisation was similar for males and females between 65 and 79 years. However, females had a higher antibiotic utilisation rate than males between the ages of 80 and 94, and men of 95 years and above had a higher utilisation rate than women (Figure 3). Marquez et al.’s study support this finding that after age 65, men have higher innate and pro-inflammatory activity and lower adaptive activity than women [33].

## Limitations

One of the major limitations of dispensing data is the lack of information on antibiotics’ indications and appropriateness. Therefore, adherence to antimicrobial therapy by the patients is not captured and accounted for when using such datasets. We used data for dispensing claims for funded antibiotics made by community pharmacies only, thus possibly missing data on unfunded antibiotics prescribed to the study population during the study period. Additionally, the Pharmaceutical Collections database does not include information about medicines dispensed in hospitals [34], so antibiotic exposure may have been underestimated if older adults received the drugs during hospitalisation. Our post-intervention observation period was relatively short (from January 2018 to December 2019) and therefore might not reflect the true impact of the intervention. Other factors besides the antimicrobial action plan may have played a significant role in the huge drop in antibiotic utilisation rates between 2018 and 2019. For example, there was already an annual 4.6% drop in antibiotic utilisation rate three years before the launch of the antimicrobial intervention plan [28].

## Strengths

The reimbursement system in New Zealand captures greater than 95% of the prescription coverage of older adults’ census population, strengthening our study findings’ validity. This study also acts as a proxy measure of adherence to national antibiotic prescribing policies and stewardship guidelines among older adults in New Zealand. In addition, the validity of our findings was strengthened by using the ARIMA models to perform the interrupted time series analysis to investigate the impact of the antimicrobial resistance action plan on antibiotic utilisation. Previous studies have demonstrated the utility and reliability of ARIMA models for evaluating population-level health interventions [18].

## Conclusion

This study provides valuable information on the consumption of systemic antibiotics among the 65 years or above sub-population of New Zealand. It has also shown the effectiveness of the antimicrobial resistance action plan introduced by the government of New Zealand to mitigate antibiotic resistance by reducing inappropriate antimicrobial utilisation. The ITS analysis has demonstrated that this intervention hugely impacted the antibiotic utilisation rate among older adults. As a result, there was a significant reduction in the overall utilisation of systematic antibiotics between 2005 and 2019 among older adults in New Zealand. However, there were some age and sex differences in the consumption rates of antibiotics among the studied population. Further studies are required to determine whether the reduced consumption rates of antibiotics are associated with reduced rates of antibiotic-associated adverse drug events such as acute kidney injury and haematological abnormalities.

## Data Availability

Data availability: The data is owned by the Analytical Services, Ministry of Health, New Zealand, so supporting data is not available.

## Ethics statement

The Departmental Research and Ethics Officer at the University of Bath assessed the ethical implications of research activity (EIRA) and approved this study on behalf of the University of Bath’s Research, Integrity, and Ethics committee. Approval number EIRA 1-4048.

## Acknowledgement

The authors thank the Analytical Services, Ministry of Health of New Zealand, for supplying the data extracted from the Pharms database.

## Author contributions

TC (Bath) serves as the first author of the manuscript. He led all the stages of the development of this manuscript while PN supervised. TC (Otago), TC (Bath), and PN contributed to statistical analysis, interpretation of results, and manuscript revision. SR and HJ also contributed to interpreting the results and manuscript revision. All authors approved the final manuscript.

## Data availability

The data is owned by the Analytical Services, Ministry of Health, New Zealand, so supporting data is not available

